# Estimating COVID-19 cases and deaths prevented by non-pharmaceutical interventions, and the impact of individual actions: a retrospective model-based analysis

**DOI:** 10.1101/2021.03.26.21254421

**Authors:** Kathyrn R Fair, Vadim A Karatayev, Madhur Anand, Chris T Bauch

## Abstract

Simulation models from the early COVID-19 pandemic highlighted the urgency of applying non-pharmaceutical interventions (NPIs), but had limited empirical data. Here we use data from 2020-2021 to retrospectively model the impact of NPIs in Ontario, Canada. Our model represents age groups and census divisions in Ontario, and is parameterised with epidemiological, testing, demographic, travel, and mobility data. The model captures how individuals adopt NPIs in response to reported cases. We compare a scenario representing NPIs introduced within Ontario (closures of workplaces/schools, reopening of schools/workplaces with NPIs in place, individual-level NPI adherence) to counterfactual scenarios wherein alternative strategies (e.g. no closures, reliance on individual NPI adherence) are adopted to ascertain the extent to which NPIs reduced cases and deaths. Combined school/workplace closure and individual NPI adoption reduced the number of deaths in the best-case scenario for the case fatality rate (CFR) from 178548 [CI: 171845, 185298] to 3190 [CI: 3095, 3290] in the Spring 2020 wave. In the Fall 2020/Winter 2021 wave, the introduction of NPIs in workplaces/schools reduced the number of deaths from 20183 [CI: 19296, 21057] to 4102 [CI: 4075, 4131]. Deaths were several times higher in the worst-case CFR scenario. Each additional 9−16 (resp. 285−578) individuals who adopted NPIs in the first wave prevented one additional infection (resp., death). Our results show that the adoption of NPIs prevented a public health catastrophe. A less comprehensive approach, employing only closures or individual-level NPI adherence, would have resulted in a large number of cases and deaths.

## Introduction

Non-pharmaceutical interventions (NPIs) such as school and workplace closure, limiting group sizes in gatherings, hand-washing, mask use, and physical distancing are essential for pandemic mitigation in the absence of a vaccine [1]. Scalable NPIs, in particular, are measures that can be taken up by the entire population if containment strategies have failed [2]. These measures have been applied extensively during the 2019 coronavirus disease (COVID-19) pandemic to reduce severe outcomes [3]. Given the extensive social and economic consequences of the COVID-19 pandemic, there is significant value in assessing how many cases, hospitalizations, and deaths were prevented by pandemic mitigation measures that relied upon scalable NPIs.

Assessments of NPI efficacy sometimes rely upon comparing health outcomes in countries that did not implement certain NPIs, to those that did [4]. However, it may be difficult to control for confounding factors in cross-country comparisons such as differing social and economic circumstances. Another approach is to monitor outcomes longitudinally in a given population as they respond to a timeline of changing NPIs [5].

Empirical approaches to predicting the number of COVID-19 cases in the absence of interventions are difficult or impossible since, in every country, governments implemented control measures and/or the population responded to the presence of the virus. Even in the case of Sweden, whose government adopted a *de facto* herd immunity strategy [6], the population exhibited enormous reductions in mobility in March and April 2020 [7]. Here, simulation models can be useful tools for estimating the number of cases in the absence of interventions, and for exploring other questions concerning SARS-CoV-2 transmission and COVID-19 disease burden [8–18]. Models developed during the pandemic’s early stages made projections for such scenarios, but required rational assumptions about crucial parameter values in the absence of empirical data specific to COVID-19 [9, 16, 18].

We adopt a retrospective approach, fitting a simulation model to empirical data from March 2020 to February 2021 to estimate how many COVID-19 cases and deaths would have occurred in the province of Ontario, Canada in the absence of NPIs. After fitting the model, we relax the parameters relating to NPIs to predict what might have happened in their absence, or in the presence of only a subset of NPIs. The model includes the census area and age structure of Ontario, as well as travel between census areas. Moreover, the model accounts for population behavioural responses to pandemic waves: without volitional population uptake of NPIs, “flattening the curve” may not have been possible [17].

## Methods

### Model overview

To capture the social-epidemiological dynamics of severe acute respiratory syndrome coronavirus 2 (SARS-CoV-2) transmission and COVID-19 cases, we developed a stochastic compartmental model incorporating age and spatial structure (Figure 1). Transmission dynamics in the population of each census region are described by a Susceptible (S), Exposed (E), Pre-symptomatic and infectious (P), Symptomatic and infectious (I), Asymptomatic and infectious (A), Removed (R) natural history. Populations in different regions are connected through commuter travel. Transmission is reduced through school and/or workplace closure and infection control efforts in those settings, under direction from public health authorities. However, transmission is also reduced outside of school and work settings as a result of volitional efforts by individuals to adopt NPIs, including measures such as physical distancing, hand-washing, and mask wearing (Supplementary information, Figure S2). This occurs in proportion to the daily incidence of reported cases. Transmission rates are region-specific to account for regional differences in contact patterns due to population density and other factors, and were also modified by seasonality in transmission. Age classes varies in their relative susceptibility. Age-specific testing rates increase over time from initially low levels in March 2020 to a constant level (with the date this is attained varying by age class).

**Figure 1.**
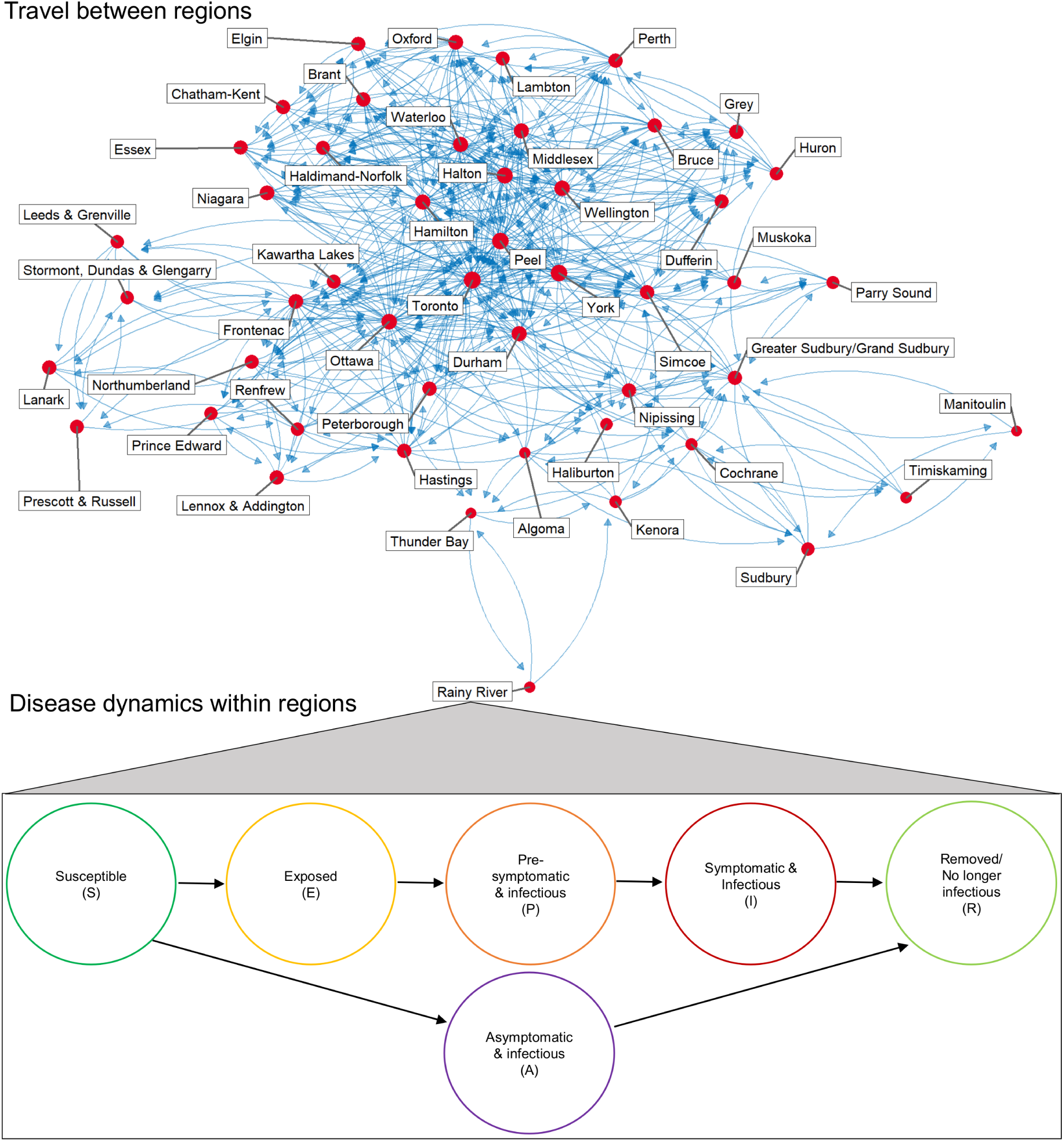
Schematic representation of transmission model. Note that the epidemiological compartments were stratified by age as well as location (details in Supplementary methods).

Using Ontario data, we estimated deaths resulting from COVID-19 under best-case and worst-case scenarios for the crude case fatality ratio (CFR). In the best-case scenario we assume that CFR computed from the historical for the first and second waves also applies in counterfactual scenarios where the case incidence was much higher due to relaxing NPIs. In the worst case, we extrapolate the observed empirical relationship between case incidence and CFR to consider the possibility that the CFR increases with case numbers [19], due to increased strain on the healthcare system [20] (Supplementary information, Figure S9).

Epidemiological [21–23], testing [21–23], demographic [24], travel [25], and mobility data [7] for Ontario were used to parameterise the model. We employed a 2-stage non-linear optimization process to fit cases by age class at the provincial level, and total cases at the Public Health Unit (PHU) level [26–28]. The first stage used a global algorithm, with the results of that fitting input as the initial values for the second-stage local optimization. This allowed us to estimate the baseline transmission rate, as well as how it responded to school/workplace closure, and how many individuals adhered to NPIs in response to reported case incidence.

Full details on the model structure and parameterisation appear in the Supplementary methods. All fitting, simulations, analysis, and visualization are performed in Rstudio (Version 1.2.5019) using R (Version 4.0.3) [29, 30]. Code and data used for parameter fitting and simulations are available in a GitHub repository (https://github.com/k3fair/COVID-19-ON-model).

### Scenarios and outcomes analyzed

We generated model outputs for reported COVID-19 cases and deaths over three time periods. The first time period covers the first wave from 10 March 2020 to 15 August 2020. The second time period from 12 June 2020 to Feb 28 2021 covers Ontario’s reopening during the first wave and the subsequent second wave. These periods are studied separately because the first and second waves differed considerably in terms of their epidemiology, disease burden, and interventions. These two time periods were analyzed retrospectively: the empirical data from these time periods were used to fit the model.

In the first time period, Ontario implemented school and workplace closure, and a significant proportion of the population adhered to recommended NPIs. For the first wave, we projected what might have happened under three counterfactual scenarios: (1) school/workplace closures were enacted but no individuals adhered to any other NPIs, (2) school/workplace closures were not enacted but individuals adhered to other NPIs in proportion to reported case incidence, and (3) school/workplace closures were not enacted and no individuals adhered to NPIs (a “do nothing” scenario).

In the second time period, Ontario closed schools and workplaces in late 2020/early 2021, and began re-opening in February 2021, but with mandatory NPIs in place to combat transmission, such as requiring mask use in schools. We considered two counterfactual scenarios for the re-opening phase in February 2021: (1) reopening does not occur (school/workplace closures continues indefinitely), and (2) schools and workplaces are reopened without NPIs in place. Individual NPI adherence varied in response to cases in homes and other locations for all of these scenarios. We also note that all of our scenarios for the second time period incorporated the first provincial imposition of control measures in Spring 2020 followed by the first provincial re-opening in Summer 2020.

For the first time period and with reference to the average population uptake of NPIs during those periods, we also estimated how many additional individuals must adopt NPIs to prevent one additional case, or one additional death (i.e., incremental cases and death prevented by NPI uptake). These measures gauge the impact of individual-level efforts on the course of the pandemic. The numbers are calculated as an incremental quantity because the incremental effectiveness of an individual choosing to adopt NPIs depends upon how many other individuals in the population are already doing so, on account of their impact on community transmission.

## Results

### Cases and deaths prevented by NPIs in the first and second waves

Results for our three counter-factual scenarios in the first wave highlight the key role that NPIs played in limiting the spread of SARS-CoV-2, and also show how school/workplace closures interact with individual-level behaviours concerning NPIs (Figure 2). The actual number of daily reported cases peaked at 640 in Ontario in April 2020, and the modelled time series of cases follows the empirical epidemic curve (Figure 2a, inset). However, in the absence of both school/workplace closure and individual uptake of NPIs, the model predicts that daily number of reported cases would have peaked at 67000 in May 2020. Allowing for either school/workplace closure or individual uptake of NPIs reduces this peak considerably, although the peaks are still large compared to the factual (historical) scenario where both were applied.

**Figure 2.**
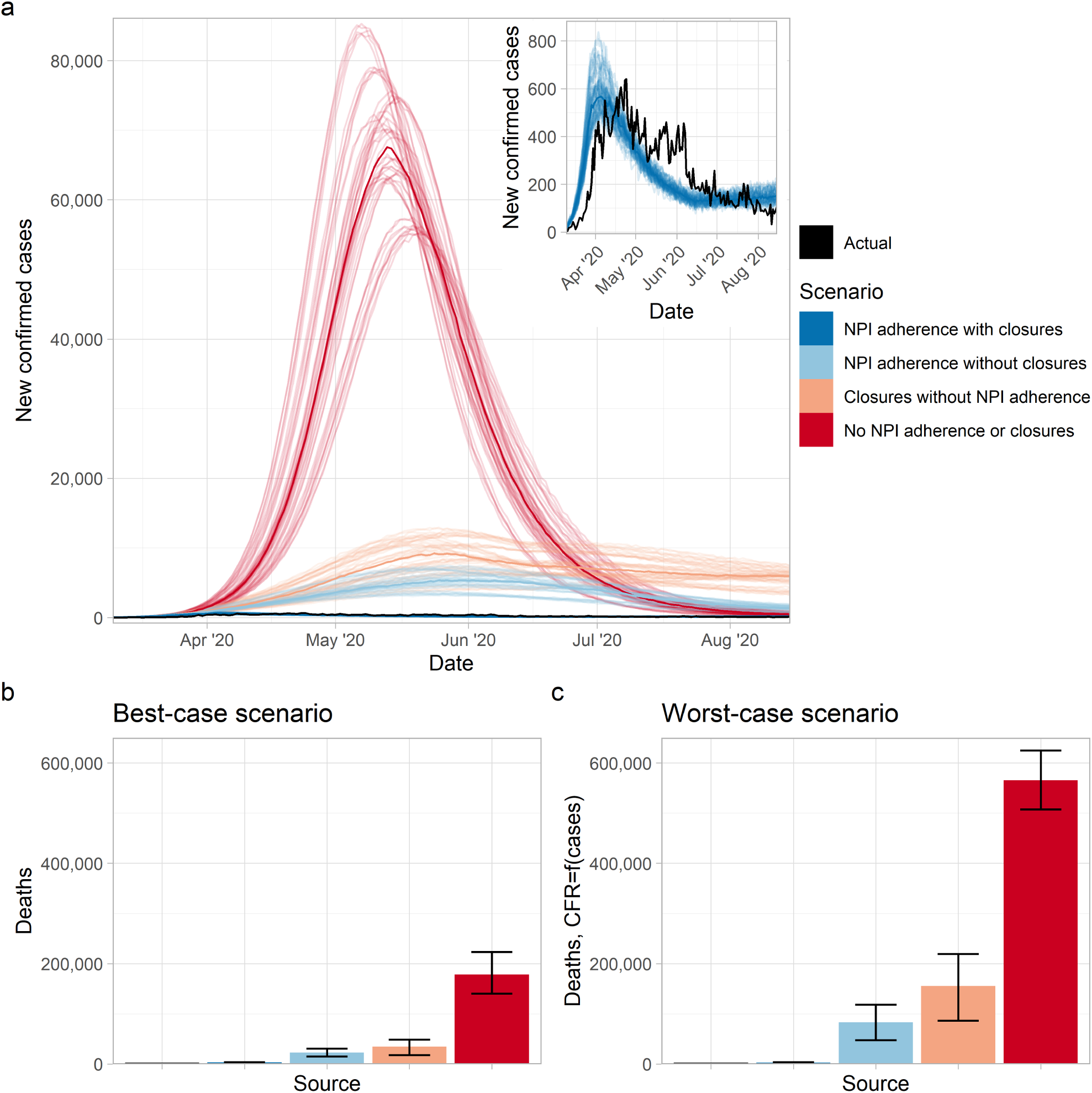
NPIs significantly reduced cases and deaths in the first wave. Figure panels show (a) new confirmed cases by day, and mean projected deaths from 10 March 2020 to 15 August 2020 in (b) the best-case scenario (values from left-right are: 2789, 3190, 22647, 34525, 178548) and (c) worst-case scenario (values from left-right are: 2789, 2493, 82727, 155388, 565218) for healthcare system functioning in a regime of very high case incidence. Transparent lines in panel (a) correspond to different stochastic realizations of model runs, with solid lines corresponding to the median value across all realizations. Error bars in panels (b,c) represent the minimal and maximal values across all stochastic realizations. For each scenario we generate 5 realizations using each of the 10 best parameter sets. Model parameter settings appear in Supplementary information, Table S1.

Under the best case scenario for the CFR, the first wave would have resulted in 178548 [CI: 171845, 185298] deaths in the absence of both school/workplace closure and individual adherence to NPIs (Figure 2b). This number greatly exceeds the 2789 deaths that actually occurred between 10 March and 15 August 2020 due to lockdown and population adoption of NPIs [31]. The worst-case scenario for deaths is even higher under the “do nothing” scenario (Figure 2c), on account of the unmanageable surge in cases causing a heightened CFR. However, applying either school/workplace closure or individual uptake of NPIs significantly reduces the number of deaths in both worst- and best-case scenarios. The reductions are greater for applying only individual-level NPI measures than for applying school/workplace closure. This is because school/workplace closure in our model only affects school-age children and working-age adults working in non-essential businesses, whereas individual adoption of NPIs in our model spans all employment sectors in all age groups.

These findings are qualitatively unchanged for the second wave, except that the difference in cases and deaths across the scenarios is not as large, since we did not evaluate a “do nothing” scenario. (Figure 3). As before, cases and deaths are considerably higher when NPI use is limited (in this case, does not occur in workplaces/schools). Both the empirical epidemic curve and the modelled epidemic curves share the feature of a relatively slow rise to a peak, followed by a relatively rapid drop afterwards (Figure 2a). This is due to the combined effect of timing of school/workplace closure, behavioural response, and seasonality in the transmission rate.

**Figure 3.**
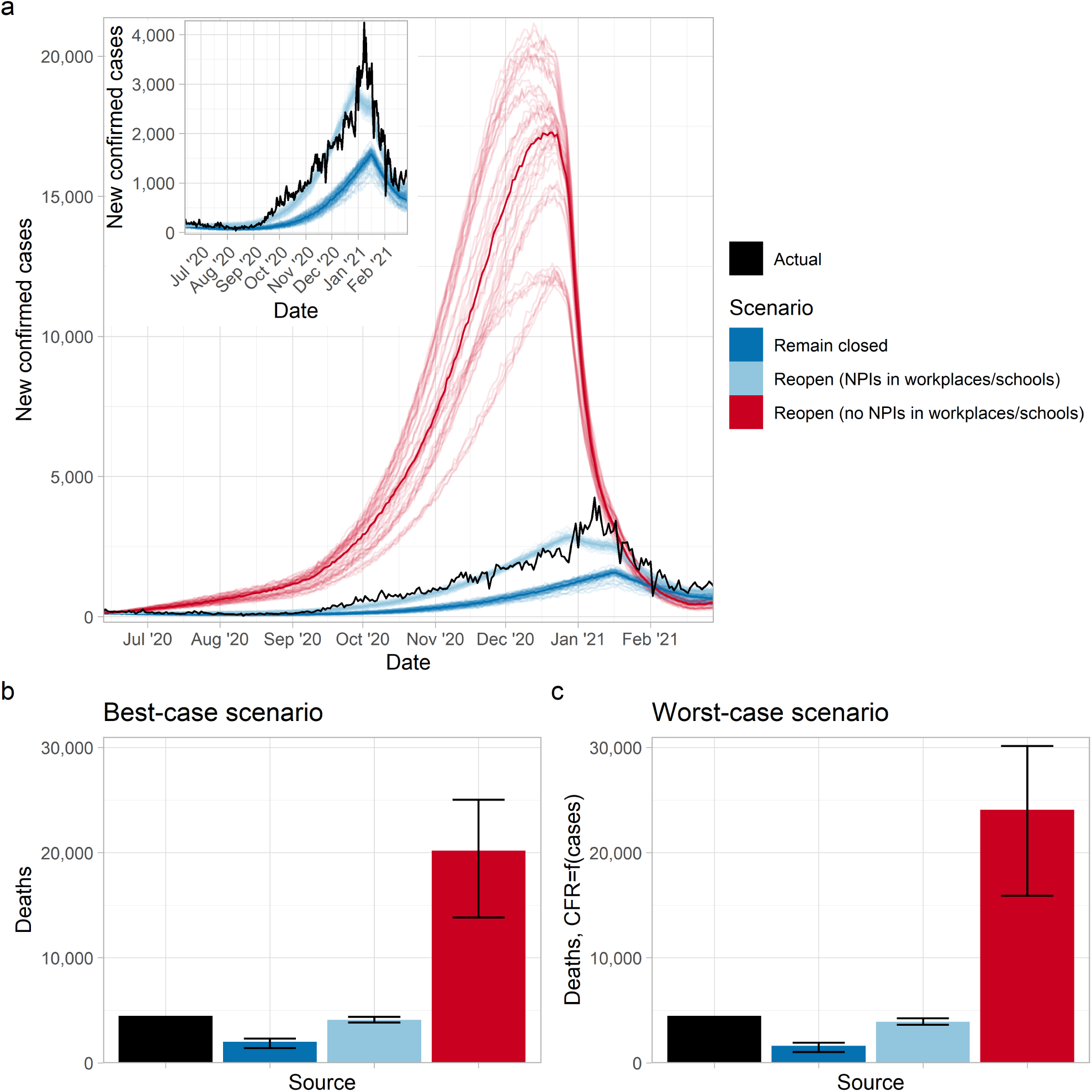
NPIs significantly reduced cases and deaths in the second wave. Figure panels show (a) new confirmed cases by day, and mean projected deaths from 12 June 2020 to 28 February 2021 in (b) the best-case scenario (values from left-right are: 4493, 2016, 4102, 20183) and (c) worst-case scenario (values from left-right are: 4493, 1636, 3908, 24060) for healthcare system functioning in a regime of very high case incidence. Transparent lines in panel (a) correspond to different stochastic realizations of model runs, with solid lines corresponding to the median value across all realizations. Error bars in panels (b,c) represent the minimal and maximal values across all stochastic realizations. For each scenario we generate 5 realizations using each of the 10 best parameter sets. Model parameter settings appear in Supplementary information, Table S1.

### Impact of individual-level efforts

We estimated how many additional individuals must adopt NPIs to prevent one additional case, and one additional death, given what percentage of the population is already adherent to NPIs. We estimated this under both best-case and worst-case scenarios for the CFR. When the percentage of the population already adherent to NPIs in within empirically valid ranges for the first wave (shaded regions in Figure 4), we estimated that every 9 to 16 individuals who adopted NPIs prevented a single SARS-CoV-2 infection. Similarly, every 285 to 578 (respectively, 184 to 474) individuals who adopted NPIs prevented a single COVID-19 death in the best-case (respectively, worst-case) scenarios.

**Figure 4.**
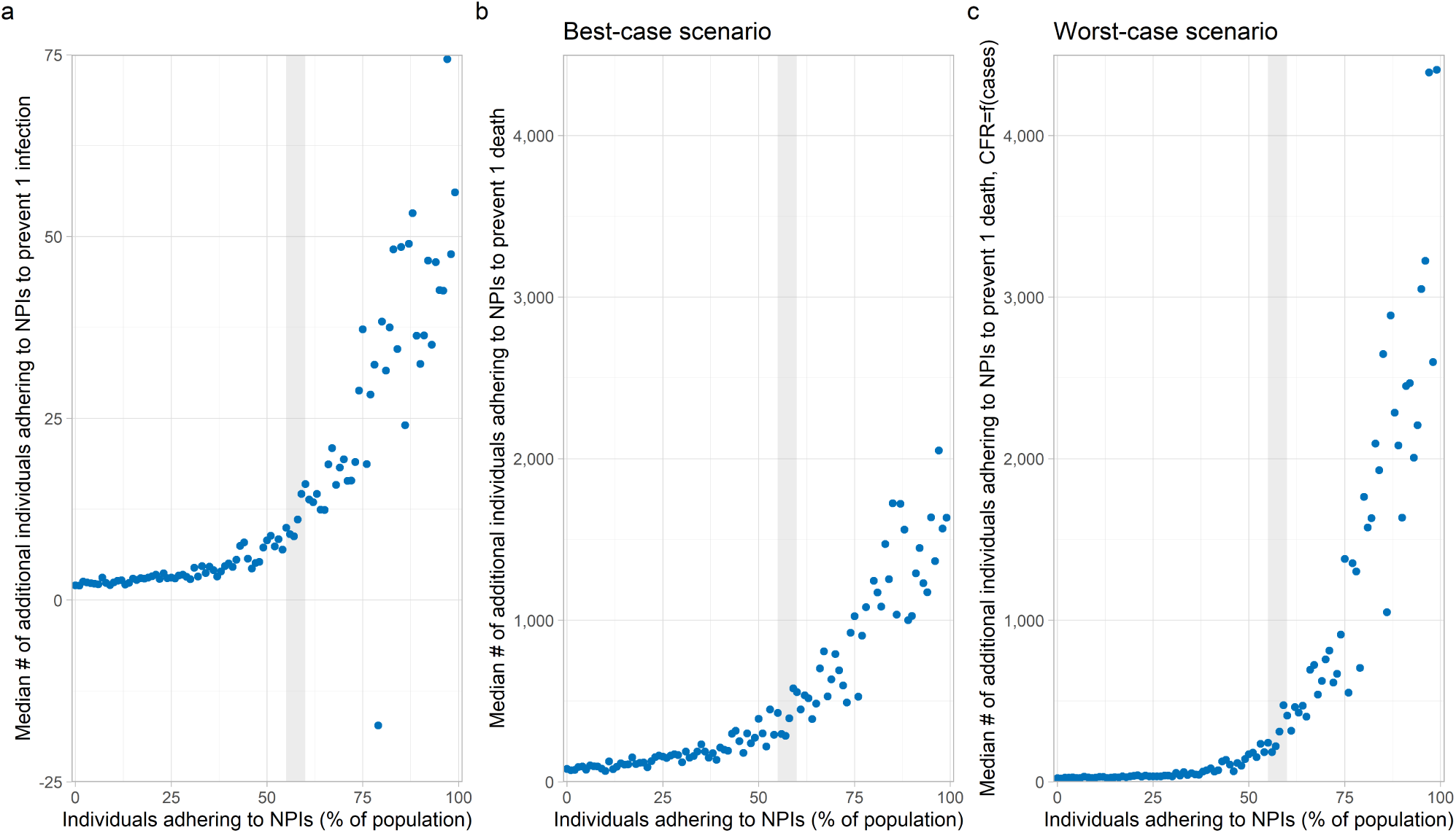
Impact of individual efforts. Figure panels show the incremental median number of individuals who needed to adopt NPIs to prevent (a) one infection, and one death under (b) the best-case scenario and (c) worst-case scenario for healthcare system functioning in a regime of very high case incidence, for the first wave (10 March to 15 August 2020). The shaded region demarcates the estimated range in the percentage of individuals adhering to NPIs over that time-period (see Supplementary information, Figure S11). We generate 5 realizations using each of the 10 best parameter sets at each value of the percentage of the population adhering to NPIs. Model parameter settings appear in Supplementary information, Table S1.

In the extreme case where a very high percentage of the population is already adherent to NPIs, the incremental number of individuals who must adhere to NPIs to prevent one case or death increases dramatically. This is expected, since high uptake of NPIs can reduce case incidence to very low levels, and thus reduce the incremental benefit of a few more individuals adopting NPIs. Similarly, in the other extreme when few individuals in the population have adopted NPIs, the incremental benefit of each additional individual who adopts NPIs is higher.

## Conclusion

This suite of simulations informs a picture of how NPIs–particularly the combination of government mandated measures such as school/workplace closure and volitional individual level actions such as physical distancing and hand-washing–strongly mitigated COVID-19 cases and deaths across both age and census area in Ontario. School/workplace closure or individual-level NPIs implemented on their own would also have reduced both cases and deaths considerably, although the absolute numbers would still have been large.

The number of deaths averted by NPIs was particularly large in the first wave. Our projection of 178548 deaths in the “do nothing” scenario for interventions and the best-case scenario for the CFR is plausible: supposing that 70% of Ontario’s 14.6 million people had been infected in a “do nothing” scenario, the adjusted CFR for Spring 2020 of 1.6% [19] would have resulted in 163520 deaths. Moreover, the actual number of deaths would likely have been much higher than suggested by our best-case scenario. The adjusted CFR of 1.6% was estimated from a population where the ICU capacity in Spring 2020 was not greatly exceeded [31]. Therefore, the adjusted CFR would have been much higher in a population contending with a massive surge in cases.

Our results on the number of individuals who must adopt NPIs to prevent one case or death increased dramatically with the percentage of the population already adhering to NPIs (Figure 4). As a result, an individual in a population where most others have already adopted NPIs has a reduced personal incentive to practice NPIs, since the number of cases (and thus their perceived infection risk) is lowered. This suggests the possibility of a free-rider effect whereby non-mitigators gain the benefits of reduced community spread without contribute to infection control [32], although social norms can curb this effect [33, 34].

Our model made several simplifying assumptions that could influence results and/or limit the conditions under which the model can be used. For instance, as our model describes community spread, we are not explicitly accounting for how transmission within congregate living settings, long-term care homes, etc. can cause case numbers to increase rapidly [31, 35, 36]. As well, our simplification of Ontario’s tiered system for NPIs at the level of individual public health units into a single aggregate “open with NPIs in place” state may lead us to over/underestimate cases at the PHU level, if the tier that PHU is in is more/less restrictive than the aggregate state.

It is well known that mathematical models can be used for forecasting purposes, but they can also be valuable for conveying insights, or for aspirational purposes. In the latter case, mathematical models can motivate the uptake of behaviours to avoid the worst-case scenarios scenarios predicted by the model. The prosocial preferences that humans often adopt toward infectious disease control [33, 34] suggest that this use of models can be effective. Early mathematical models developed during the COVID-19 pandemic showed us what might happen if we chose not to mitigate the pandemic. Our retrospective analysis that uses data from the past year confirms that we prevented a very large loss of life by our decision to take action, and that each individual person who chose to adopt NPIs helped prevent both cases and deaths.

## Data Availability

Data sets required to run simulations are available in a GitHub repository (https://github.com/k3fair/COVID-19-ON-model). Data sets generated from our analysis and simulations are available from the corresponding author upon reasonable request. All epidemiological, testing, demographic, travel, and mobility data used to parametrise the model are publicly available online.

## Author contributions

**Kathyrn R. Fair:** methodology, software, validation, formal analysis, investigation, data curation, writing - original draft preparation, writing - reviewing & editing, visualization.

**Vadim A. Karatayev:** software, validation, data curation, writing - reviewing & editing.

**Madhur Anand:** conceptualization, methodology, supervision, writing - reviewing & editing.

**Chris T. Bauch:** conceptualization, methodology, supervision, resources, writing - original draft preparation, writing - reviewing & editing.

## Acknowledgements

This research was made possible by the facilities of the Shared Hierarchical Academic Research Computing Network (SHARCNET: www.sharcnet.ca) and Compute/Calcul Canada.

## Data availability

Data sets required to run simulations are available in a GitHub repository (https://github.com/k3fair/COVID-19-ON-model). Data sets generated from our analysis and simulations are available from the corresponding author upon reasonable request. All epidemiological [21–23], testing [21–23], demographic [24], travel [25], and mobility data [7] used to parametrise the model are publicly available online.

## Code availability

Code used for parameter fitting and simulations is available in a GitHub repository (https://github.com/k3fair/COVID-19-ON-model). Code used for analysis and visualization is available from the corresponding author upon reasonable request.

