## Supplementary information for "Estimating COVID-19 cases and deaths prevented by non-pharmaceutical interventions, and the impact of individual actions: a retrospective model-based analysis"

### Supplementary methods

#### Base model

Our model, modified from [1], captures transmission dynamics for SARS-CoV-2 within Ontario, Canada. Age structure is introduced, with age classes 1-5 respectively represent 0-19 years old, 20-39 years old, 40-59 years old, 60-79 years old, and 80+ years old. The model also includes mechanisms for testing, individual NPI adherence, and the implementation of closures (of schools and workplaces).

We describe the transmission dynamics according to a $SEPAIR$ disease progression. Individuals are classed according to their epidemiological status; susceptible to infection ($S$), exposed i.e. infected but not yet infectious ($E$), pre-symptomatic and infectious ($P$), asymptomatic i.e. infectious without ever developing symptoms ($A$), symptomatic and infectious ($I$), or removed i.e. no longer infectious ($R$). We also incorporate testing for the virus, with individuals classed as having either a known ($K$) or unknown ($U$) infection status. Testing occurs for symptomatic, pre-symptomatic, and symptomatic individuals and the outcome of these tests becomes known with some daily probability.

Ontario's population is distributed across 49 census divisions, hereafter referred to as regions [2]. Within our model, each region contains a population with the same size as the census division it represents. As the region-specific population data is from the 2016 census, we use the 2020 Q4 population estimate for Ontario to scale up all region population sizes [3]. Each day within the simulation begins with every individual within age class $i$ in region $j$ travelling to region $k$ for the day, with some probability $\nu_i m_{jk}$. Individuals who travel to another region experience any transmission events in the region they are visiting. At the end of the day, these individuals return to region $j$.

The travel matrix, $M = [m_{jk}]$ contains empirical data on the frequency of travel between regions [2]. We use a matrix $M^* = [2m_{jk}]$ as input to our model, since the data collected considers only individuals aged 15 and up who commute from their home to their place of work and thus excludes travel by unemployed individuals and those under age 15, as well as travel undertaken for other reasons (shopping, athletics, social events, etc.). We supplement $M^*$ with age-specific travel rates, assuming that young/old individuals travel less frequently than working-age individuals Individuals in age classes 1,4,5 (ages 0-19, 60+) reduce their travel by a factor of $\nu_i = \nu_0$, with $\nu_i = 0$ for age classes 2,3 (ages 20-39, 40-59).

As 81% of reported COVID-19 cases are mild, we assume infected individuals reduce their travel by a factor $r = 0.19$ [4]. Additionally, individuals who test positive for COVID-19 reduce their travel by a factor $\eta = 0.8$ [5, 6], so the combined reduction in travel for an infected individual in age classes 1, 4 or 5 who has tested positive is $(1 - \nu_i)(1 - r)(1 - \eta)$. When a region closes its schools, the rate of travel to that region is reduced by a factor $\epsilon_s$, with a similar reduction, $\epsilon_w$, when its workplaces are closed. When both are closed, the combined reduction in travel is $(1 - \epsilon_s)(1 - \epsilon_w)$. Similarly, if a region has its schools or workplaces open, but with NPIs in place, travel is reduced by a factor of $\delta_s\epsilon_s$ and of $\delta_w\epsilon_w$ respectively. These parameters

($\epsilon_w,\ \epsilon_s, \delta_w, \delta_s$) are explained in further detail below, as they are also used to describe the efficacy of NPIs in reducing contacts in schools and workplaces.

As noted in [1], there is a risk of overestimating the effect of travel, since we assume that all individuals within an age class have an equal likelihood of travelling to another region on any given day. Within real-world populations, the same individuals will tend to travel consistently (e.g. will travel frequently, to the same region, and interact with the same group of contacts). Despite this, it is preferable to err on the side of over-estimation, as travel can substantially impact spread of SARS-CoV-2 through the importation of the virus to regions with few or no cases.

Each individual, $i$, has a state, $\{D_i^j, T_i^j\}$ based on their age class, $j \in \{1, 2, 3, 4, 5\}$, epidemiological status, $D_i^j \in \{S^j, E^j, P^j, A^j, I^j, R^j\}$, and testing status, $T_i^j \in \{U^j, K^j\}$. An individual with age class $j$, epidemiological status $D^j$, and either testing status is $\{D_i^j, \cdot\}$, similarly for $\{\cdot, T_i^j\}$. Within region $k$ on day $t$, $P_{t,k}^{D^jT^j}$ is the number of individuals with age class $j$ and state $\{D_i^j, T_i^j\}$, $P_{t,k}^{DT}$ is the number of individuals across all age classes with state $\{D_i, T_i\}$, and $P_{t,k}$ is the total population size.

Every day, states for individuals with age class $j$ in region $k$ are updated according to the following steps:

1. Exposure: With probability $\lambda_{j,k}(t)$, $\{S^j, \cdot\}$ individuals are exposed to SARS-CoV-2 and shift to $\{E^j, \cdot\}$.

2. Onset of infectious period: With probability $(1 - \pi)\alpha$, $\{E^j, \cdot\}$ individuals become pre-symptomatic and infectious and transition to $\{P^j, \cdot\}$, or alternatively (with probability $\pi\alpha$) become asymptomatic and infectious and transition to $\{A^j, \cdot\}$.

3. Onset of symptoms: With probability $\sigma$, individuals in $\{P^j, \cdot\}$ become symptomatic and shift to $\{I^j, \cdot\}$.

4. Testing: With probability $\tau_{k,I^j}$, individuals in $\{I^j, U^j\}$ are tested and shift to $\{I^j, K^j\}$, similarly for $\tau_{k,P^j}$ and $\tau_{k,A^j}$.

5. Removal: With probability $\rho$, individuals in $\{I^j, \cdot\}$ and $\{A^j, \cdot\}$ cease to be infectious and are removed to $\{R^j, \cdot\}$.

During the onset of the infectious period, newly infected individuals are assigned as super-spreaders with probability $s = 0.2$ [7]. Super-spreaders are denoted with the subscript $s$, while non-super-spreaders are denoted by $ns$, creating sub-divisions within $P^j$, $A^j$ and $I^j$ given by $P_s^j$, $P_{ns}^j$, $A_s^j$, $A_{ns}^j$, $I_s^j$, $I_{ns}^j$. The probability of a super-spreader infecting others is $(1 - s)/s$ higher than the probability of a non-super-spreader doing so. Several other factors also impact the infection probability. First, the individual's contacts; both those in schools and workplaces (limited by closures) and those in homes and other locations (unaffected by closures). Second; the prevalence of the virus within the population. Third; the effectiveness of closures and individual adherence to NPIs in reducing transmission.

Individuals risk contracting or transmitting SARS-CoV-2 when they interact with others. Four possible locations for interactions are assumed; schools ($s$), workplaces ($w$), homes ($h$), and other ($o$). For each location, a contact matrix $N^l = [n^l_{ij}]$, where $l \in \{s, w, h, o\}$, contains information on age-stratified contact frequencies. Each $n^l_{ij}$ indicates the relative frequency with which age class $i$ has contacts in age class $j$, normalized using the the highest total number of contacts (across all locations) for a single age class. Matrices (Figure S1) are generated by aggregating Canada-specific data from [8]. The 75-80 age class in [8] is used as a proxy for our 80+ age class. When aggregating, we weight the data from each 5-year age class by the proportion of Ontario's population that falls within that age-range [9].

Closures reduce contacts in schools ($s$), workplaces ($w$). NPIs introduced to combat SARS-CoV-2 transmission in workplaces and schools as they reopen will also reduce contacts, to a lesser extent. $C^l_k(t)$ controls the measures in place for workplaces ($l = w$) or schools ($l = s$) in region $k$:

$$C^l_k(t) = \begin{cases} 0 \text{ if } l \text{ are completely open,} \\ \delta_l \epsilon_l \text{ if } l \text{ are open with NPIs in place,} \\ \epsilon_l \text{ if } l \text{ are closed.} \end{cases} \tag{1}$$

Parameters $\epsilon_{w,s}$ represent the efficacy of closures in workplaces, and schools. Additionally, $\delta_{w,s} < 1$ control how effective NPIs in workplaces and schools are, in comparison to a closure.

All remaining contacts are assumed to occur at home ($h$) or in other locations ($o$), where contacts may be reduced through individual adherence to NPIs. The maximum efficacy of NPIs in homes is denoted by $\epsilon_h$, similarly for other locations by $\epsilon_o$. In region $k$, the level of individual adherence to NPIs in these locations, $\chi_k(t)$, is a function of perceived risk [10, 11], based on the prevalence of confirmed cases within the region's population, and of the presence/absence of stay-at-home orders in the region, according to

$$\chi_k(t) = 1 - e^{-\left(\omega(t)\left(P^+_{t,k}/P_{t,k}\right) + L(t)\right)} \tag{2}$$

where $P^{AK}_{t,k} + P^{IK}_{t,k} = P^+_{t,k}$ indicates the number of active confirmed cases in the region and

$$L(t) = \begin{cases} 0 \text{ if no stay-at-home orders,} \\ L_0 \text{ if stay-at-home orders in place,} \end{cases} \tag{3}$$

with $L_0$ capturing the extent to which stay-at-home orders impact NPI adherence. The risk perception coefficient, $\omega(t)$, changes over time, beginning to decrease during the second wave according to

$$\omega(t) = \begin{cases} \omega_0 \text{ if } t < t_d \\ \omega_0 e^{-\zeta(t - t_d)} \text{ if } t \geq t_d \end{cases} \tag{4}$$

where $\omega_0$ is the base risk perception coefficient, $t_d$ corresponds to August 15, 2020 and $\zeta$ controls the decay of $\omega(t)$ over time.

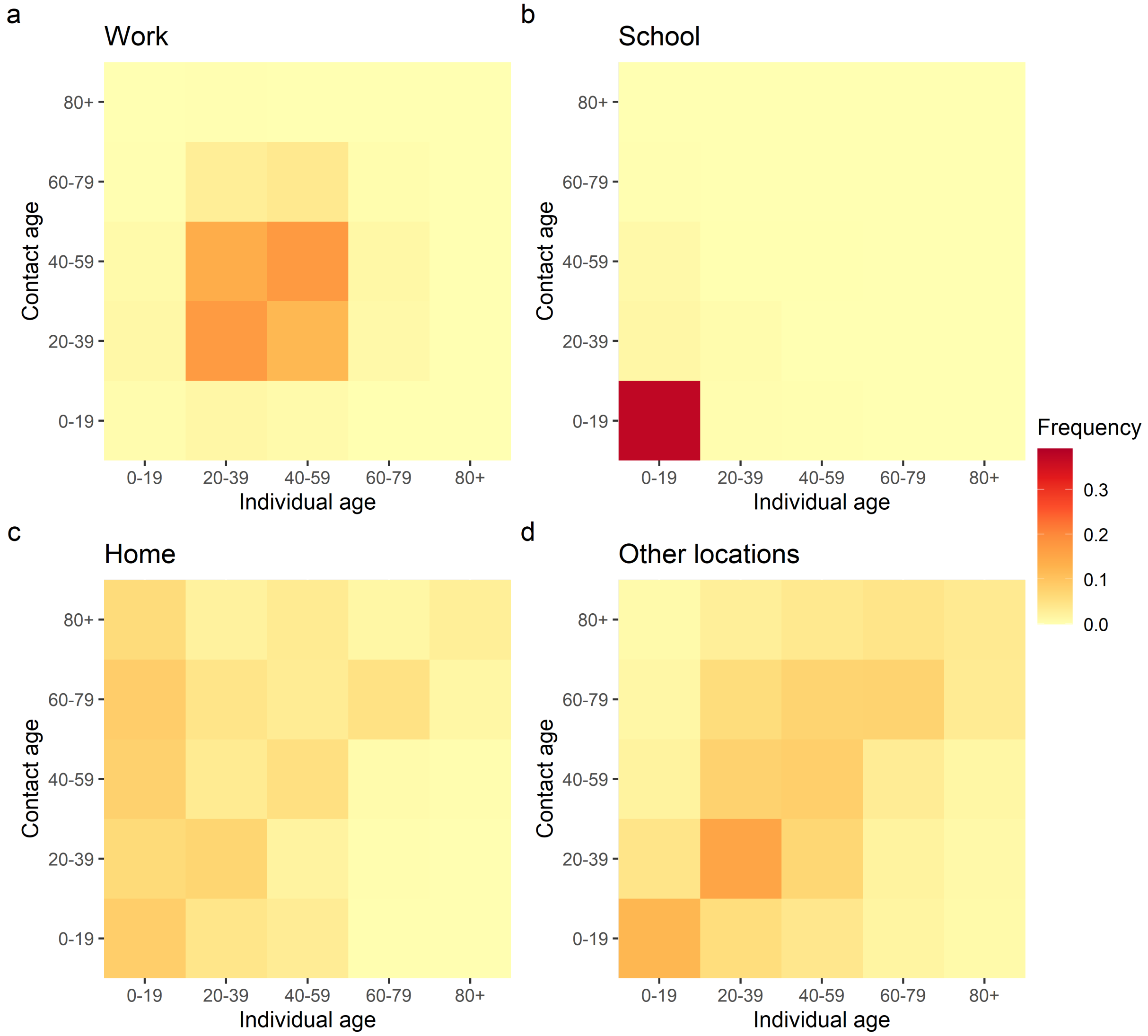


**Figure S1. Relative frequency of contacts between individuals within different age classes.** Matrices are aggregated from [8] to align with our age-classes. Labels above subpanels indicate which location the subpanel corresponds to. Frequencies are normalized on the highest total number of contacts for an age-class across all locations.

Allowing $\omega(t)$ to change over time captures both the idea that perceived risk changes as new information about SARS-CoV-2 and COVID-19 becomes available, and that, though not explicitly modelled, the accumulating costs (economic, social, etc.) associated with NPI adherence may lead individuals to reduce their level of adherence [12, 13]. Ontario-specific data suggests it is plausible to assume these factors are influencing NPI adherence As time passes, the extent to which mobility in retail and recreation locations [14] (our proxy for NPI adherence) decreases when active cases are high is lessened (Figure S2a), in the absence of stay-at-home orders in one or more PHU (i.e. prior to 14 January 2021). Once stay-at-home orders have been introduced, reductions in mobility for a given level of active cases are larger.

Additionally, we estimate a provincial weighted mean value for $\omega(t)$ (Figure S2b) using Ontario-specific data, which begins to decay around mid-August 2020 (we choose this as 15 August 2020 for simplicity). The increase in the estimated $\omega(t)$ value occurring in late December 2020 (prior to the stay-at-home orders) is likely caused by a combination of changing mobility patterns over the winter holiday period, and the declaration of a province-wide shutdown on 26 December 2020. We emphasize that this is a rough estimate of $\omega(t)$, as there is not a one-to-one relationship between PHU and the regions reported in the mobility data, and include it only to illustrate the assumed trend over time.

Accounting for the effects of closures and individual NPI adherence, the fraction of contacts in age class $j$ which remain for individuals in age class $i$ at time $t$ in region $k$ is:

$$F_{ij,k}(t) = n_{ij}^{w}\left(1 - C_k^w(t)\right) + n_{ij}^{s}\left(1 - C_k^s(t)\right) + n_{ij}^{h}\left(1 - \epsilon_h \chi_k(t)\right) + n_{ij}^{o}\left(1 - \epsilon_o \chi_k(t)\right). \tag{5}$$

The transmission probability for non-super-spreaders $(\{P_{ns}, \cdot\}, \{A_{ns}, \cdot\}, \{I_{ns}, \cdot\})$ is $\beta_{D_{ns}} = \beta_0^D$, while for super-spreaders $(\{P_s, \cdot\}, \{A_s, \cdot\}, \{I_s, \cdot\})$ we set $\beta_{D_s} = \beta_0^D(1-s)/s$. Individuals who test positive for COVID-19 reduce their contacts by a fraction $\eta$, such that while $f_{T=U}$ $= 1$, $f_{T=K} = 1 - \eta$, where $\eta = 0.8$ [5, 6]. The daily probability of a susceptible individual in age class $i$ becoming infected in region $k$, stated as 1 less the probability of the individual not becoming infected, is:

$$\lambda_{i,k}(t) = 1 - \prod_{D^j, T^j} \left[1 - F_{ij,k}(t) f_T \beta_{i,k}^D(t)\right]^{P_{t,k}^{*D^j T^j}} \tag{6}$$

where $\beta_{i,k}^D(t)$ is the probability of an individual in region $k$ with epidemiological state $D$ transmitting SARS-CoV-2 to a susceptible individual in age class $i$, at time-step $t$. This probability is:

$$\beta_{i,k}^D = \xi_k \gamma_i \beta_0^D \left[1 + B\cos\left(\frac{2\pi}{365}(t+\phi)\right)\right] \tag{7}$$

where $\gamma_i$ is an age-specific susceptibility coefficient, $\xi_k$ is a PHU-specific transmission modifier, and seasonality is controlled by $B$ and $\phi$ so transmission probability peaks sometime in fall/winter and attains its lowest value in spring/summer [15]. Due to epidemiological data being reported at the PHU level $\xi_k$ values are PHU-specific not region-specific and all regions within the same PHU will have identical $\xi_k$ values. The starred notation in $P_{t,k}^{*D^j T^j}$ indicates the number of individuals with state $\{D^j, T^j\}$ in region $k$ at time $t$ after adjusting for travel.

Based on examination of Ontario-specific data on tests completed per day [16], we assume symptomatic testing probability for individuals in age class $i$ changes over time according to

$$\tau_{I_i}(t) = \begin{cases} \tau_{I_0} \text{ if } t < t_{n=50} \\ \tau_{I_{\max}} - (\tau_{I_{\max}} - \tau_{I_0}) e^{-\psi_i (t - t_{n=50})} \text{ if } t \geq t_{n=50} \end{cases} \tag{8}$$

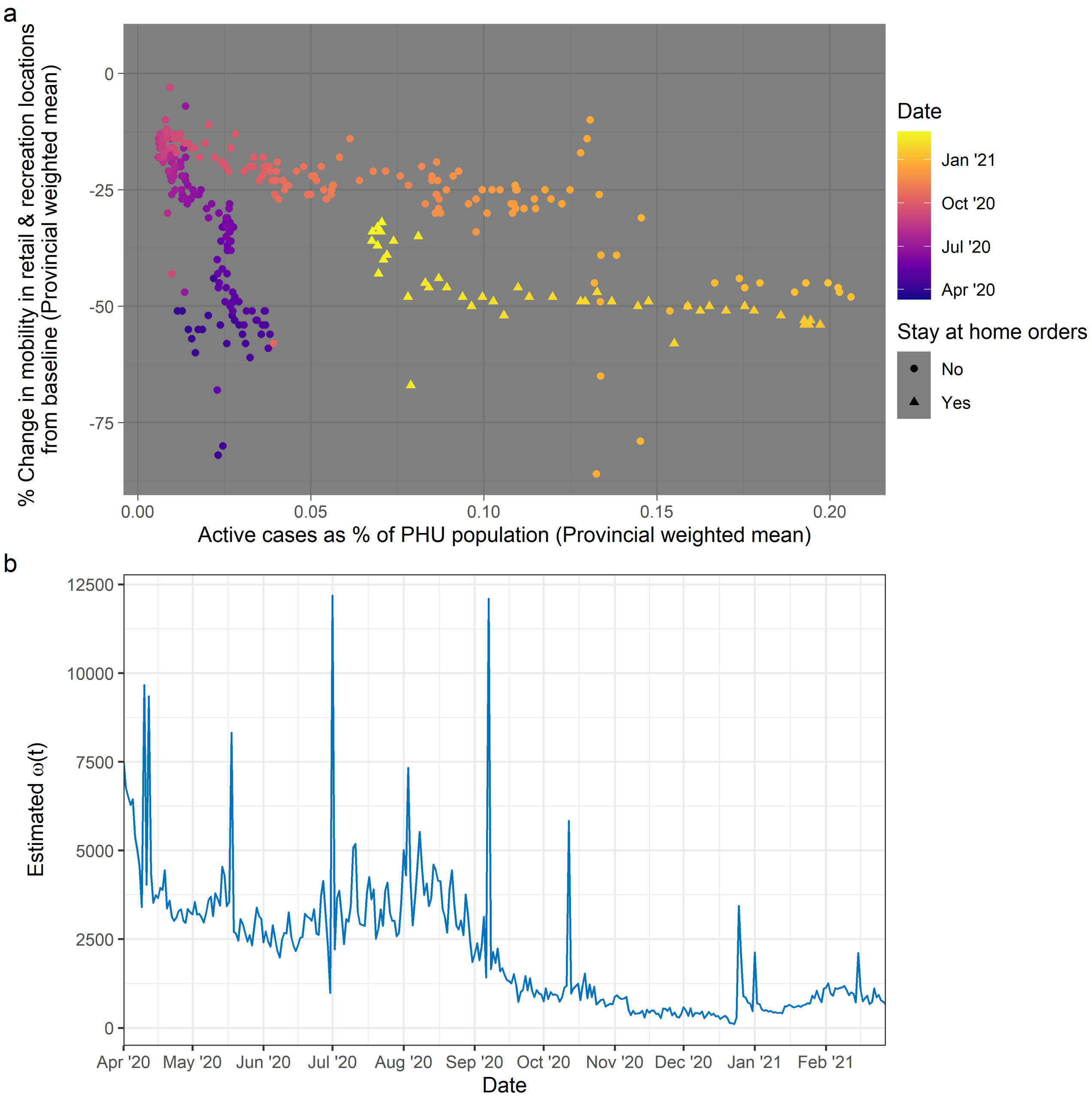


**Figure S2. Trends in NPI adherence.** a) Relationship between active case level and changes to mobility in retail and recreation locations [14] (our proxy for NPI adherence). "Stay-at-home orders" indicates whether at least 1 PHU within Ontario is under stay-at-home orders on a given date. b) Estimate of a provincial weighted mean $\omega(t)$ value, taken over $\omega(t)$ values calculated at the regional level.

where we have initial, $\tau_{I_0}$, and maximum $\tau_{I_{\max}}$ testing probabilities that hold across all age classes. For each age class, $i$, we fit values for the parameter, $\psi_i$, controlling the rate at which the testing probability increases over time. Age-specific testing rates are assumed, as factors such as targeted testing in long-term care homes and schools, and the severity of symptoms in different age groups can influence the likelihood that a person of a given age is tested [17–19].

We assume that pre-symptomatic and asymptomatic individuals experience a lower testing probability than symptomatic individuals, as provincial testing guidelines limit asymptomatic testing to high risk individuals and those in groups targeted for testing [17]. We set $\tau_{P_{0_i}} = \tau_{A_{0_i}} = 0 \ \forall \ i$. The testing probability for pre-symptomatic individuals in age class $i$ is:

$$\tau_{P_i}(t) = \begin{cases} 0 \text{ if } t < t_{n=50}, \\ \tau_{P_{\max}} \left(1 - e^{-\psi_i(t - t_{n=50})}\right) \text{ if } t \geq t_{n=50}. \end{cases} \tag{9}$$

Similarly, for asymptomatic individuals we have a testing probability:

$$\tau_{A_i}(t) = \begin{cases} 0 \text{ if } t < t_{n=50}, \\ \tau_{A_{\max}} \left(1 - e^{-\psi_i(t - t_{n=50})}\right) \text{ if } t \geq t_{n=50}. \end{cases} \tag{10}$$

For simplicity we assume $\tau_{P_{\max}} = \tau_{A_{\max}} = \kappa\tau_{I_{\max}}$, where $\kappa \in (0, 1)$ and thus

$$\tau_{P_i}(t) = \tau_{A_i}(t) = \begin{cases} 0 \text{ if } t < t_{n=50} \\ \kappa\tau_{I_{\max}} \left(1 - e^{-\psi_i(t - t_{n=50})}\right) \text{ if } t \geq t_{n=50} \end{cases} \tag{11}$$

To implement key events within simulations, and compare our results to empirical data, we use the day the 50th case was detected in Ontario ($t_{n\geq50}$), 10 March 2020 [20], as our reference point. All time-steps (days) within simulations are measured in relation to $t_{n\geq50}$. Ontario declared a state of emergency on 17 March 2020, and we assume no NPI adherence occurred prior to this ($\omega = 0$ for $t - t_{n\geq50} < 7$). Within the model, the measures (closures, reopeninngs) for each PHU apply to all regions within that PHU.

Schools were closed for March Break as of 14 March 2020 and remained closed for the rest of the 2019-2020 school year. We use 8 September 2020 as our school reopening date. Workplaces were closed on 25 March 2020 and we treat the day the majority of Ontario entered Phase 2 of the reopening plan (12 June 2020) as our workplace reopening date. In both cases, reopenings occur with NPIs in place to combat SARS-CoV-2 transmission. Schools closed for the Winter Break on 21 December 2020. A province-wide shutdown came into effect on 26 December 2020 and was upgraded to a stay-at-home order on 14 January 2021, wherein Ontario's population was required to remain at home except for essential trips. During February/March 2021 these orders began to lift, on 8 February 2021 schools reopened in all PHU except for Toronto, Peel, and York (where schools reopened on 16 February 2021). On 10 February 2021 workplaces reopened in a small number of PHU (Hastings Prince Edward, Kingston, Frontenac and Lennox & Addington, and Renfrew County). For the majority of PHU, workplaces reopened on 16

February 2021, with York held back until 22 February 2021 and Toronto, Peel, and North Bay - Parry Sound until 8 March 2021. We assume that schools in PHU were closed from 21 December 2020 until their February 2021 reopening dates, and likewise that workplaces were closed from 26 December 2021 until their February/March 2021 reopening dates. Stay-at-home orders were in effect from 14 January 2021 until the day that the workplaces in a PHU reopened in February/March 2021.

#### Parameterisation

We set $\beta_0^{P,A} = 0.5\beta_0^I$, based on our assumed period of infectiousness and data indicating that 44% of SARS-CoV-2 shedding occurs prior to the onset of symptoms [21]. All parameters not obtained from the literature are estimated by fitting modelled (1) time series of newly confirmed cases in each age class (number of individuals in each age class entering $\{\cdot, K\}$ states each day) to data on daily confirmed cases (by reporting date and age) at the provincial level [20]; (2) time series of PHU-level totals for newly confirmed cases across all age classes (aggregated from the region-level model output) to data on daily confirmed cases (by reporting date) at the PHU level [22]; (3) underascertainment ratio (ratio of total cases to confirmed positive cases) at the provincial level to a empirically estimated underascertainment ratio of 8.76 for the United States [23]; and (4) individual NPI adherence, $\xi_k(t)$, to a real-world proxy - the change in mobility trends for retail and recreational locations [14]. The percent change in mobility for these locations (from baseline) is treated as indicative of the percent adherence to NPIs. All parameters are described in Table S1 and all epidemiological [16, 20, 22], testing [16, 20, 22], demographic [9], travel [2], and mobility data [14] used to parameterise the model are publicly available online.

We employ a 2-stage fitting process, first using a global non-linear optimization algorithm (Improved Stochastic Ranking Evolution Strategy [24]), and then feeding the results of that process into a second local optimizer algorithm (Constrained Optimization BY Linear Approximations [25]) to refine the solution. Optimization processes are implemented using the Nloptr package for R [26]. During both stages, the algorithms attempt to minimize the value of a cost function which incorporates all of the fitting criteria outlined above. As our global algorithm is stochastic and thus different runs of the fitting process may result in different solutions, we run the process 1000 times. Parameter distributions for the 10 best parameter sets (i.e. those with the lowest cost function value at the end of the fitting) are shown in Figure S3, Figure S4, and Figure S5. Output from simulations run using these parameter sets is shown in Figure S6, Figure S7, and Figure S8.

**Table S1. Model parameters.** Values are drawn from empirical data where possible, with remaining parameters fitted based on data from [14, 16, 20, 22, 23]. Fitted values for all parameters are shown in Figure S3, Figure S4, and Figure S5.

| Parameter | Description | Baseline value | Source |
|---|---|---|---|
| $\tau_{I_0}$ | Initial daily symptomatic testing probability | fitted | |
| $\tau_{I_{\max}}$ | Final daily symptomatic testing probability | fitted | |
| $\psi_i$ | Parameter controlling rate of increase over time in testing probabilities for age class $i$ | fitted | |
| $\kappa$ | Coefficient controlling daily pre-symptomatic and asymptomatic testing probabilities | fitted | |
| $\alpha$ | Probability of $E \to A$ transition | 0.4/day | [27, 28] |
| $\sigma$ | Probability of $A \to I$ transition | 0.4/day | [27, 28] |
| $\rho$ | Probability of $I \to R$ and $A \to R$ transitions | 0.67/day | [27, 28] |
| $s$ | Superspreader parameter | 0.2 | [7] |
| $\pi$ | Proportion of individuals who are asymptomatic | 0.2 | [29] |
| $\omega_0$ | Base risk perception proportionality constant | fitted | |
| $\zeta$ | Decay constant controlling reduction in $\omega(t)$ over time | fitted | |
| $B$ | Coefficient controlling impact of seasonality on transmission | fitted | |
| $\phi$ | Seasonality phase | fitted | |
| $L_0$ | Impact of the presence of stay at home orders on NPI adherence | fitted | |
| $\epsilon_{w,s}$ | Efficacy of closures in workplaces ($w$) and schools ($s$) | fitted | |
| $\epsilon_{h,o}$ | Maximum efficacy of NPI adherence in homes ($h$) and other locations ($o$) | fitted | |
| $\delta_{w,s}$ | Coefficients controlling efficacy NPIs in workplaces ($w$) and schools ($s$) | | |
| $\beta_0^{P,A}$ | Daily transmission probability, pre-symptomatic ($P$) and asymptomatic ($A$) | $\beta_0^{P,A} = 0.5\beta_0^I$ | [21, 23, 30] |
| $\beta_0^I$ | Daily transmission probability, symptomatic | fitted | [21, 23, 30] |
| $\gamma_i$ | Coefficient controlling rate of transmission to individuals in age class $i$ | fitted | |
| $\xi_k$ | Coefficient controlling rate of transmission to individuals in region $k$ | fitted | |
| $\eta$ | Adherence to self-isolation | 0.8 | [5, 6] |
| $r$ | Reduction to travel rate if symptomatic | 0.19 | [4] |
| $m_{jk}$ | Travel matrix | see Methods | [2] |
| $n_{ij}$ | Contact matrix | see Methods | [8] |
| $\nu_0$ | Coefficient for age-specific travel rate modifier | fitted | |
| $\mu_0$ | Coefficient for age-specific testing rate modifier | fitted | |

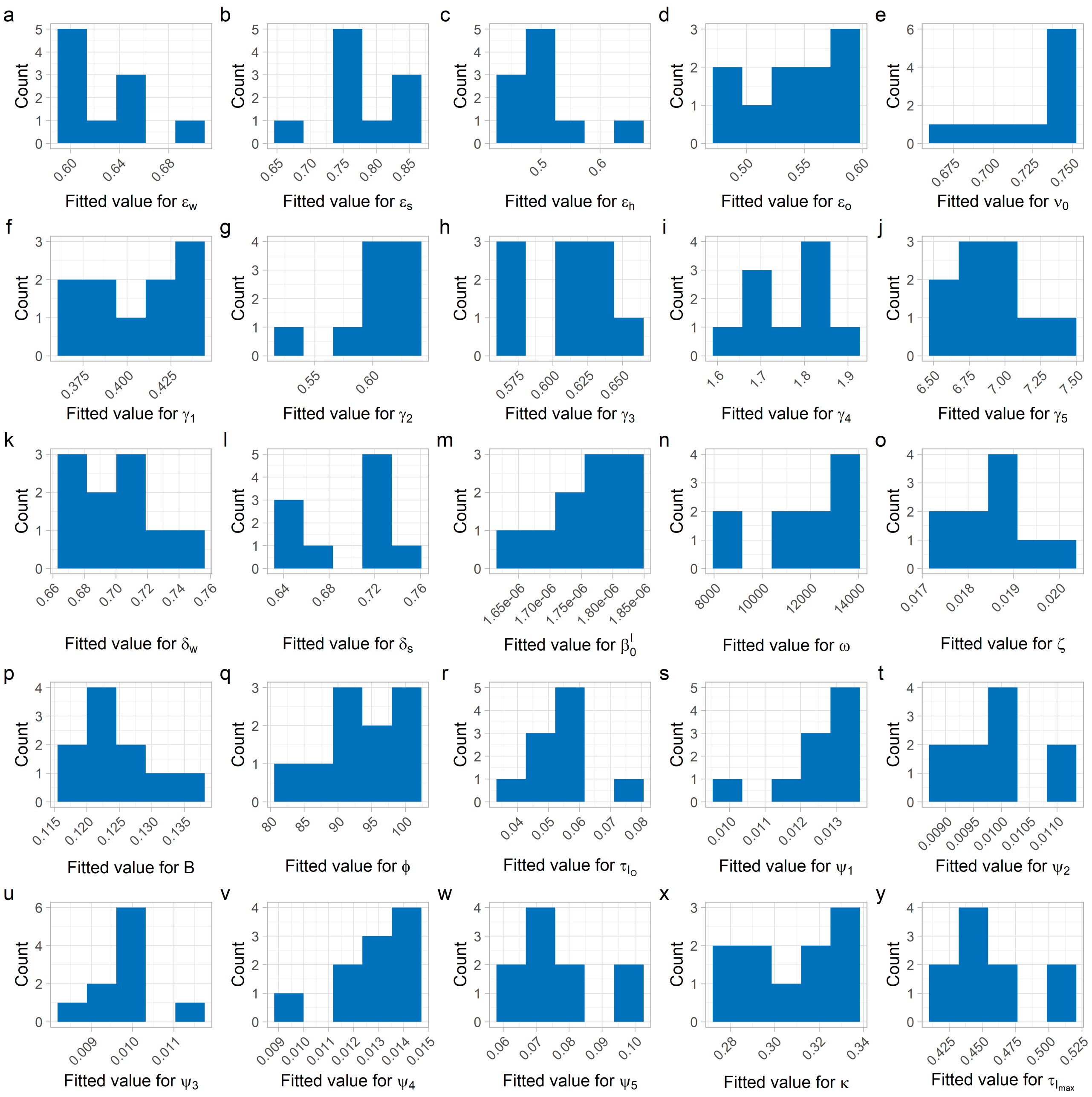


**Figure S3. Fitted parameters.** Distributions show values for individual parameters in our 10 best parameter sets.

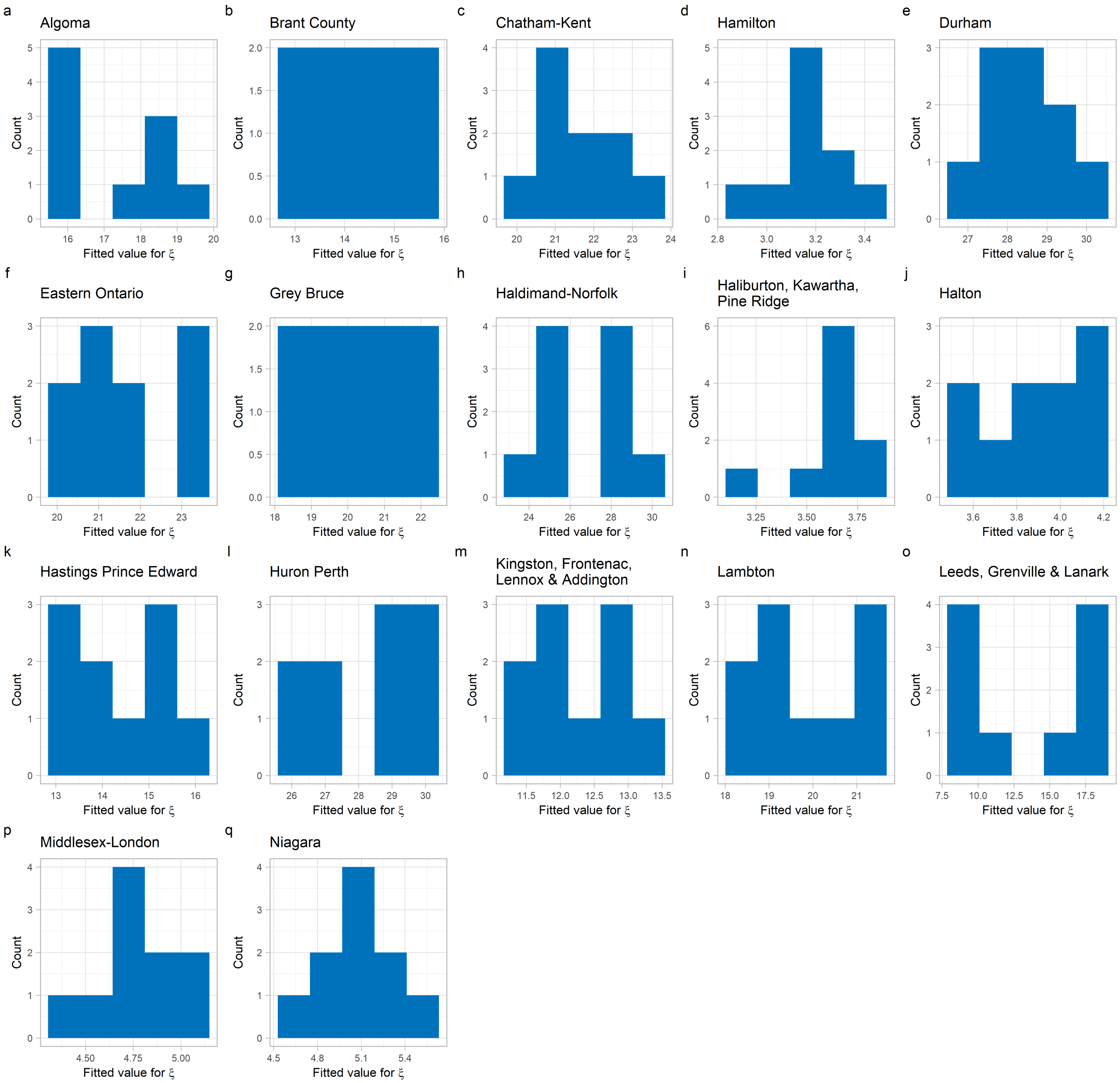


**Figure S4. Fitted PHU-specific transmission coefficients.** Distributions show values for PHU-specific transmission coefficients from our 10 best parameter sets. Each PHU-specific coefficient is applied to all regions within that PHU. The transmission coefficients for the rest of Ontario's 34 PHU are shown in Figure S5.

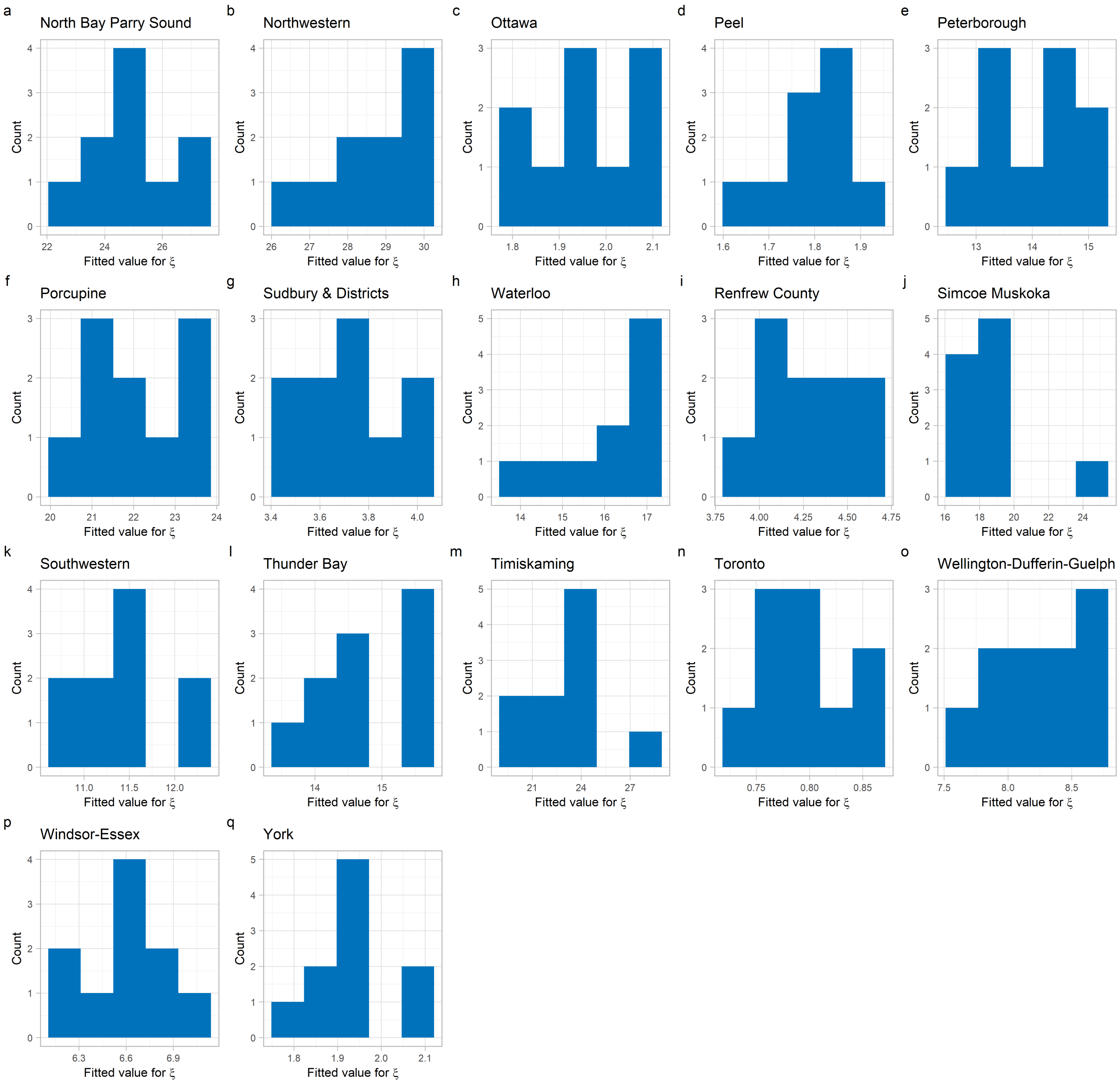


**Figure S5. Fitted PHU-specific transmission coefficients.** Distributions show values for PHU-specific transmission coefficients from our 10 best parameter sets. Each PHU-specific coefficient is applied to all regions within that PHU. The transmission coefficients for the rest of Ontario's 34 PHU are shown in Figure S4.

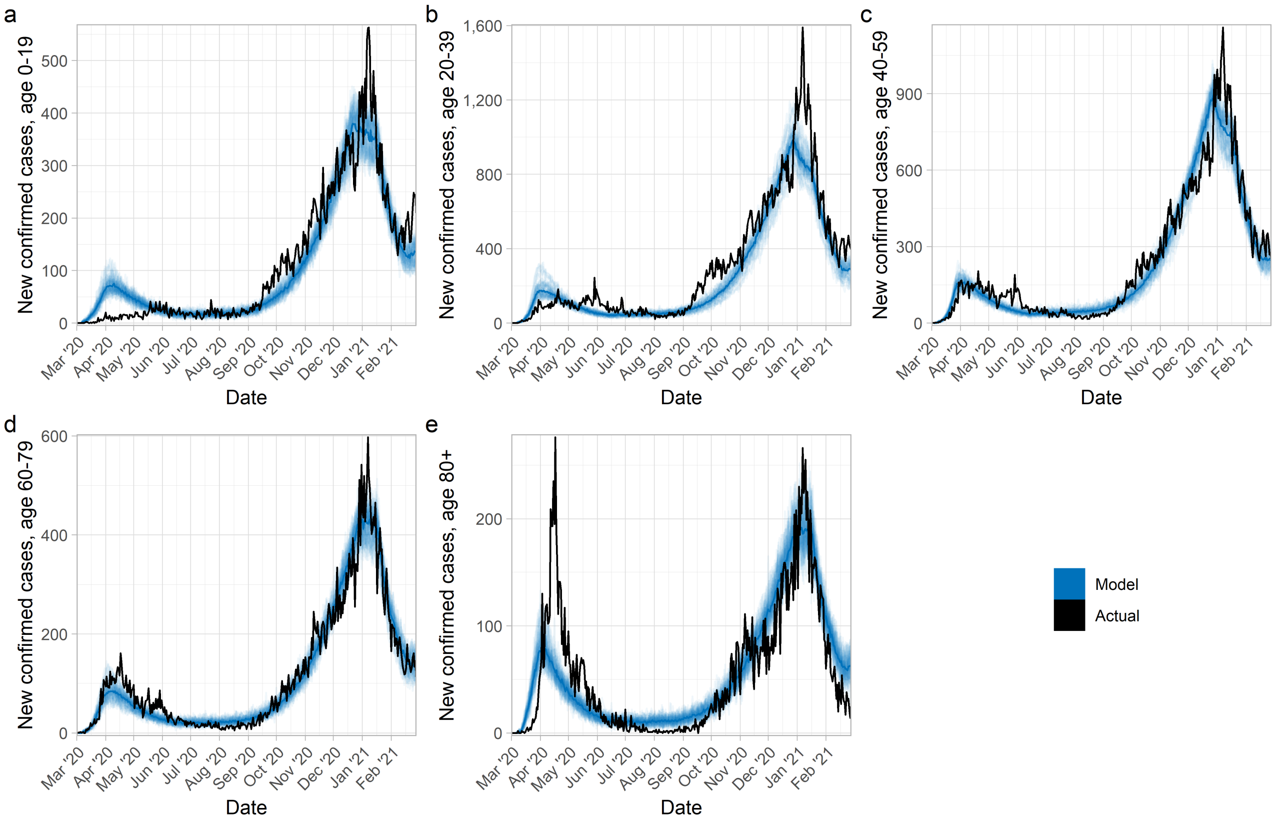


**Figure S6. Cases by age-class at the provincial level.** Transparent lines correspond to different stochastic realizations of model runs, with solid lines corresponding to the median across all realizations. We generate 5 realizations using each of the 10 best parameter sets. Model parameter settings appear in Table 1.

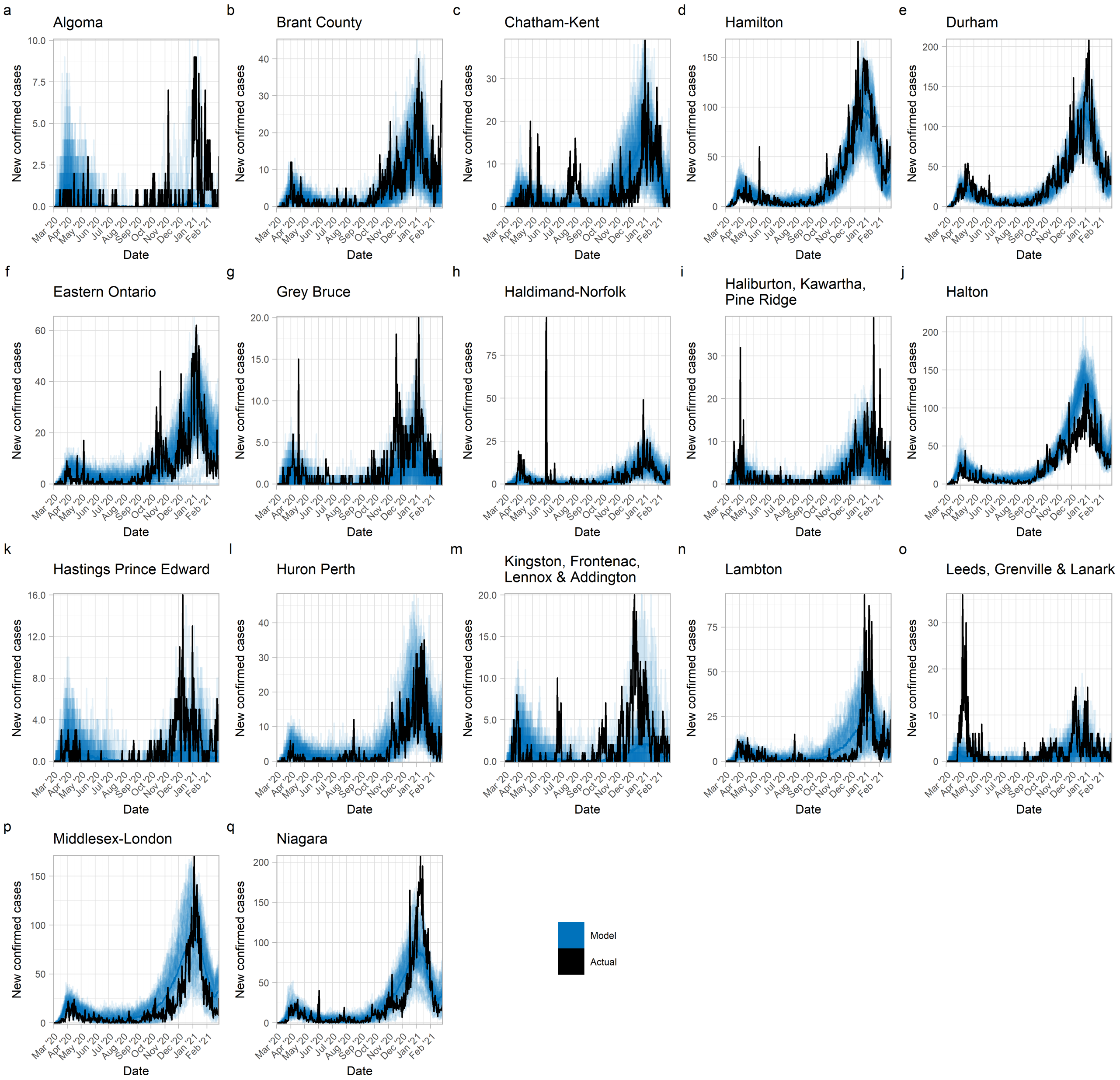


**Figure S7. Total cases across all ages at the PHU level.** Transparent lines correspond to different stochastic realizations of model runs, with solid lines corresponding to the median across all realizations. The rest of Ontario's 34 PHU are shown in Figure S8. We generate 5 realizations using each of the 10 best parameter sets. Model parameter settings appear in Table 1.

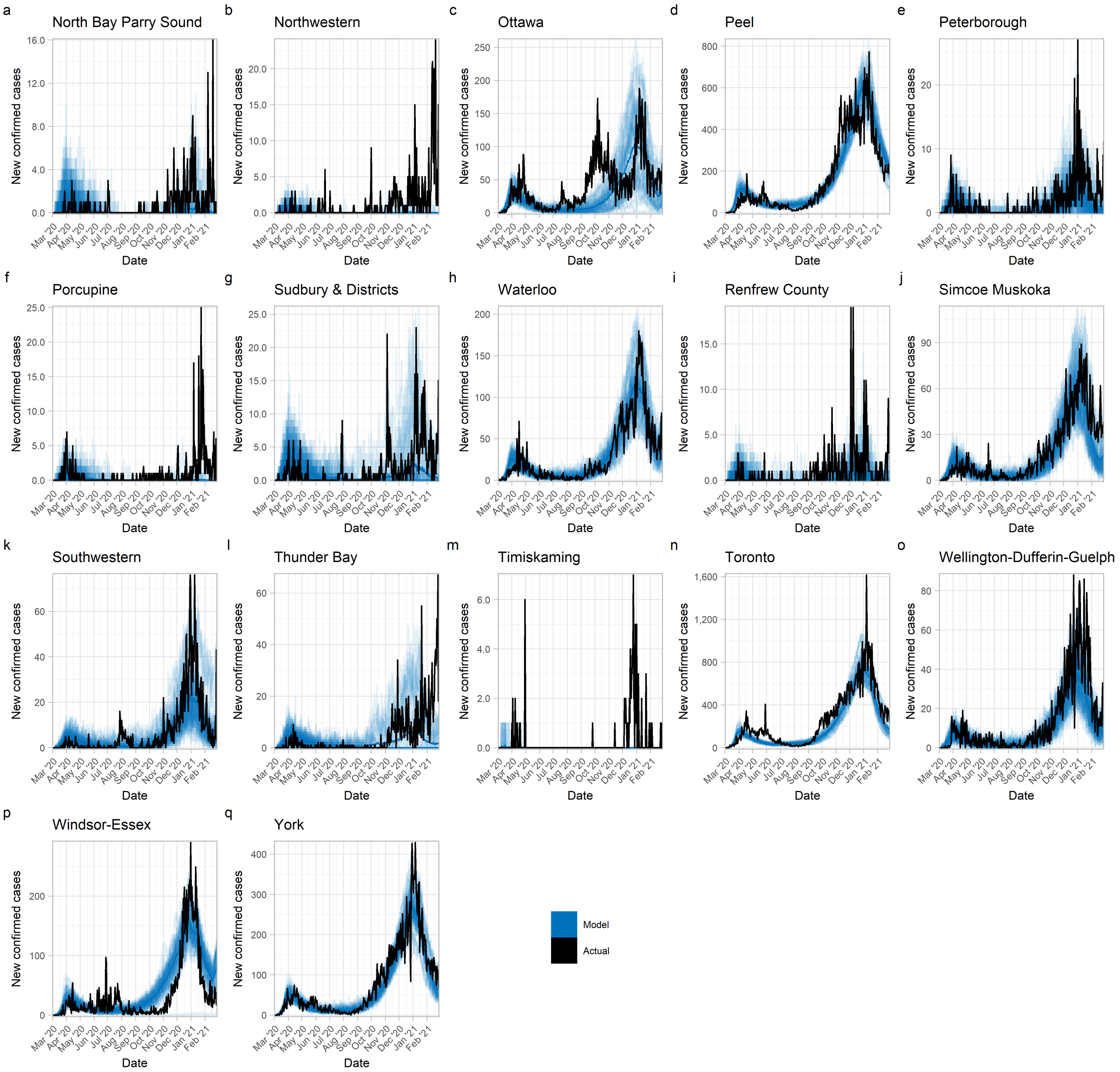


**Figure S8. Total cases across all ages at the PHU level.** Transparent lines correspond to different stochastic realizations of model runs, with solid lines corresponding to the median across all realizations. The rest of Ontario's 34 PHU are shown in Figure S7. We generate 5 realizations using each of the 10 best parameter sets. Model parameter settings appear in Table 1.

#### Case fatality ratio

To estimate deaths resulting from COVID-19, we consider 2 scenarios for the crude case fatality ratio (CFR). For the best-case scenario the CFR is calculated using Ontario-specific data [16] at weekly intervals, with the crude CFR in week $t$ given by

$$(\text{crude CFR})_t = \frac{(\text{Total deaths})_t}{(\text{Total new cases})_{t-2}}. \tag{12}$$

A 2-week lag between new cases and deaths is used based on estimates of the interval between symptom onset and a case being reported in Ontario [20] and of the interval between symptom onset and death and between case reporting and death [31–33]. Here, we assume that the CFR value in week $t$ holds regardless of the number of new cases.

However, increased strain on the healthcare system [34] may mean that the CFR increases with case numbers [31]. We consider this as an alternative worst-case scenario and fit functions of the form

$$(\text{crude CFR})_t = \delta + \mu \left(1 - e^{\nu(\text{Total new cases})_{t-2}}\right) \tag{13}$$

to the crude CFR values calculated for the best-case scenario, over different periods of time. The base CFR is controlled by $\delta$, the maximum is $\delta + \mu$ and $\nu$ controls the rate at which the CFR increases with case numbers.

Based on the clustering of weekly CFR values plotted vs. weekly cases at 2-week lag (Figure S9) we identify 3 time periods (23 March 2020 to 31 May 2020, 1 June 2020 to 16 August 2020, 17 August 2020 to 28 February 2021) during which the relationship between new cases and the CFR appeared distinct. Using these groups of points we fit parameter values for Equation 13. When fitting, we assume that $\delta$ must be no smaller than the lowest calculated CFR value (approx. 0.0025), and that $\delta + \mu \leq 0.2$ based on national-level CFR values [35]. For the 23 March 2020 to 31 May 2020 period we exclude from the fitting 3 points corresponding to weeks early in the pandemic where reporting issues result in unusually high CFR values. Fitted values of $(\delta, \mu, \nu)$ for our 3 time periods are (0.01226, 0.1877, 0.0001832), (0.002500 0.1975, 0.00008455), and (0.009264, 0.1113, 0.00008747) respectively. Estimated deaths are calculated as

$$(\text{Deaths})_t = (\text{crude CFR})_t (\text{Total new cases})_{t-2}. \tag{14}$$

To evaluate the accuracy of our CFR functions, we apply them to data on cases in the province and find a reasonable level of agreement between reported deaths and deaths calculated using Equation 13 (Figure S10).

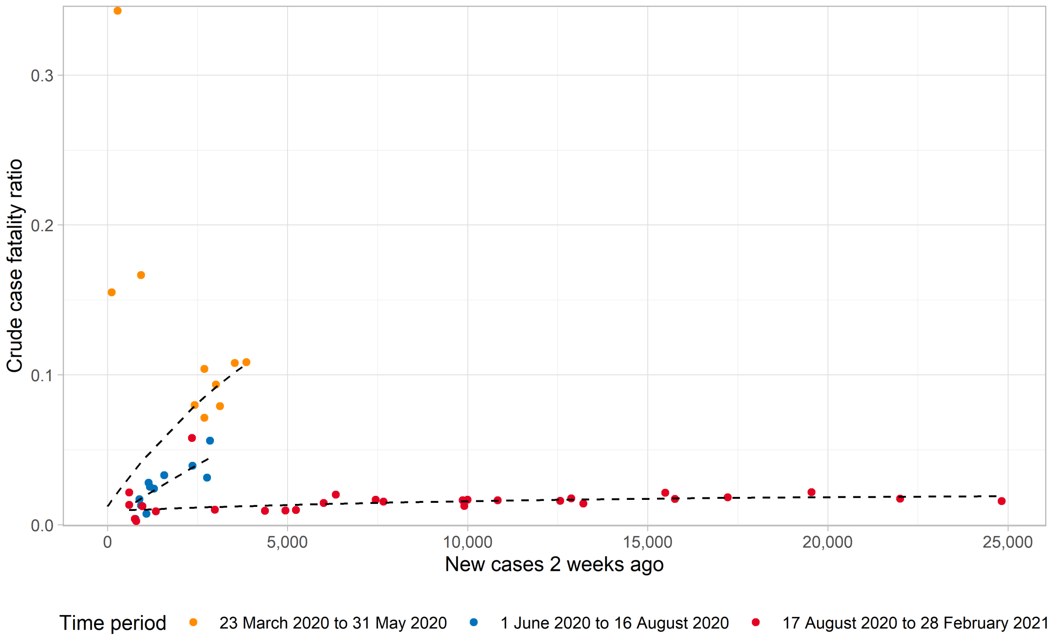


**Figure S9. Crude case fatality ratio and new cases.** Calculations based on weekly provincial totals, dashed lines indicate the crude CFR functions fit to each time period.

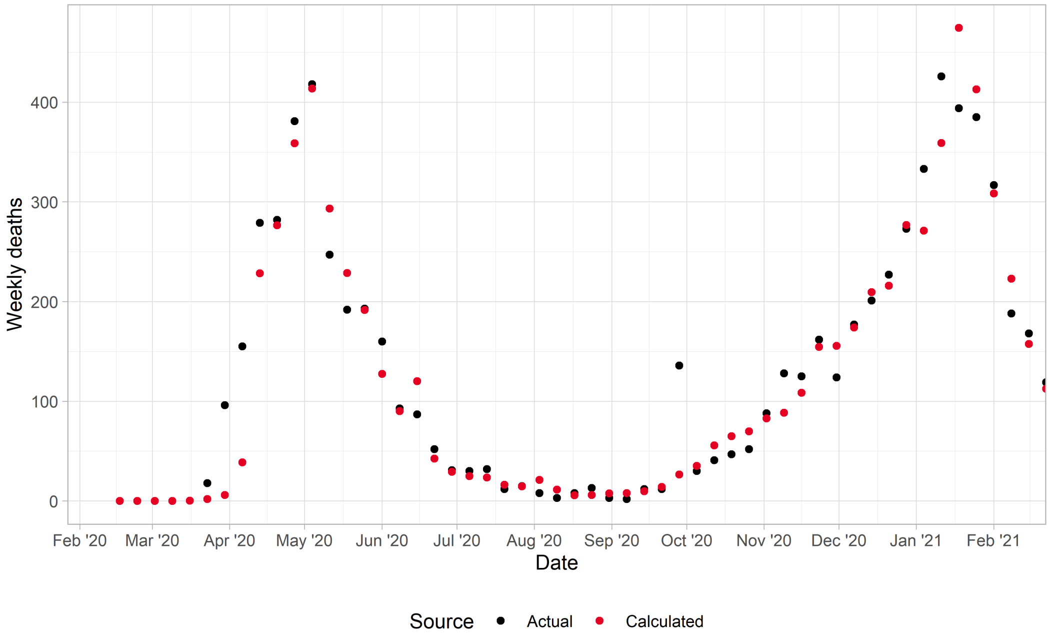


**Figure S10. Comparison between actual deaths, and deaths as calculated using crude CFR functions.** Input for CFR functions is the total number of new cases in the province 2 weeks prior.

#### Extension to individual-level NPI adherence

In our base model, Equation 2 captures, at the population level, how the proportion of the population who adhere to NPIs changes in response to case prevalence. Here, we examine NPI adherence at a more granular level. We consider a population where each individual either adheres or does not adhere to NPIs, and does not switch their behaviour during the simulation. We limit this experiment to our first time period (10 March 2020 to 15 August 2020) as over a longer time period we would expect some individuals to change their adherence.

Within this model extension some constant proportion, $x$, of the population does not adhere to NPIs (with the remaining $1-x$ adhering). This is achieved by adding a third state variable, $B_i^j$, for NPI adherence, so the state of individual $i$ in age class $j$ is $\{D_i^j, T_i^j, B_i^j\}$. Thus, $B_i^j \in \{C^j, N^j\}$ for individuals who are, respectively, adhering and not adhering to NPIs. Non-adherent individuals are seeded throughout the population, with the number of non-adherent individuals within age class $j$ in region $k$ proportional to the number of individuals within that age class in that region. The seeding of infections within the population allows for both adherent and non-adherent individuals to be amongst those initially infected.

For every interaction between a susceptible individual and an infectious individual there are 4 possibilities for NPI adherence; both parties are adherent ($CC$), neither party is adherent ($NN$), the susceptible individual is adherent but the infectious individual is not ($CN$) and vice versa ($NC$). When both parties are adherent, the proportion of contacts remaining (replacing Equation 5) is:

$$F_{ij,k}^{CC}(t) = n_{ij}^w \left(1 - C_k^w(t)\right) + n_{ij}^s \left(1 - C_k^s(t)\right) + n_{ij}^h (1 - \epsilon_h) + n_{ij}^o (1 - \epsilon_o). \tag{15}$$

If neither party is adherent the remaining fraction of contacts for a susceptible individual is:

$$F_{ij,k}^{NN}(t) = n_{ij}^w \left(1 - C_k^w(t)\right) + n_{ij}^s \left(1 - C_k^s(t)\right) + n_{ij}^h + n_{ij}^o, \tag{16}$$

with no reduction in contacts in either of the “home” or “other” locations.

When only one party is adhering to NPIs ($CN$ or $NC$) the efficacy of NPI adherence in reducing contacts depends on which party is adherent. When only the susceptible is adherent the remaining fraction of contacts is:

$$F_{ij,k}^{CN}(t) = n_{ij}^w \left(1 - C_k^w(t)\right) + n_{ij}^s \left(1 - C_k^s(t)\right) + n_{ij}^h \left(1 - \theta\epsilon_h\right) + n_{ij}^o \left(1 - \theta\epsilon_o\right) \tag{17}$$

while when only the infectious individual is adherent the remaining fraction of contacts is:

$$F_{ij,k}^{NC}(t) = n_{ij}^w \left(1 - C_k^w(t)\right) + n_{ij}^s \left(1 - C_k^s(t)\right) + n_{ij}^h \left(1 - (1-\theta)\epsilon_h\right) + n_{ij}^o \left(1 - (1-\theta)\epsilon_o\right) \tag{18}$$

where $\theta \in [0, 1]$ indicates the relative importance of the susceptible's choice to adhere with NPIs. For simplicity, we assume $\theta = 0.5$ meaning the adherence of the susceptible and infectious individual are equally important and $F_{ij,k}^{CN}(t) = F_{ij,k}^{NC}(t)$.

During the simulation, adherent susceptibles experience an infection probability:

$$\lambda_{i,k}^{C}(t) = 1 - \left( \prod_{D^j,T^j,C^j} \left[1 - F_{ij,k}^{CC}(t) f_T \beta_{i,k}^{D}\right]^{P_{t,k}^{*D^j T^j C^j}} \prod_{D^j,T^j,N^j} \left[1 - F_{ij,k}^{CN}(t) f_T \beta_{i,k}^{D}\right]^{P_{t,k}^{*D^j T^j N^j}} \right) \quad (19)$$

while non-adherent susceptibles experience an infection probability:

$$\lambda_{i,k}^{N}(t) = 1 - \left( \prod_{D^j,T^j,C^j} \left[1 - F_{ij,k}^{NC}(t) f_T \beta_{i,k}^{D}\right]^{P_{t,k}^{*D^j T^j C^j}} \prod_{D^j,T^j,N^j} \left[1 - F_{ij,k}^{NN}(t) f_T \beta_{i,k}^{D}\right]^{P_{t,k}^{*D^j T^j N^j}} \right). \quad (20)$$

### Supplementary figures

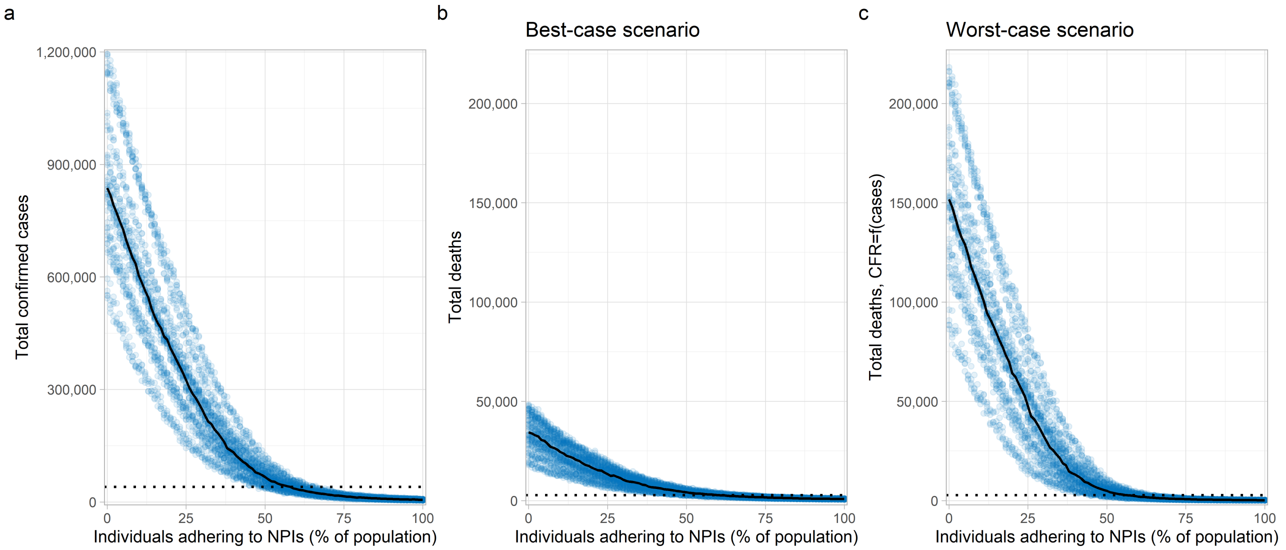


**Figure S11. Impact of individual efforts on case and death totals.** Figure panels show the total number of (a) confirmed cases and deaths under (b) the best-case scenario and (c) worst-case scenario for healthcare system functioning in a regime of very high case incidence, for the first wave (10 March to 15 August 2020). Each point indicates a realization of the model. 5 realizations are generated for each of the 10 best parameter sets at each value of the percentage of the population adhering to NPIs. Estimates of the % of the population adhering to NPIs are obtained by determining the point of intersection between actual case and death totals (horizontal dotted lines) and the corresponding median across all realizations (solid lines). Model parameter settings appear in Table 1.
